# Analysis of the early Covid-19 epidemic curve in Germany by regression models with change points

**DOI:** 10.1101/2020.10.29.20222265

**Authors:** Helmut Küchenhoff, Felix Günther, Michael Höhle, Andreas Bender

## Abstract

We analyze the Covid-19 epidemic curve from March to end of April 2020 in Germany. We use statistical models to estimate the number of cases with disease onset on a given day and use back-projection techniques to obtain the number of new infections per day. The respective time series are analyzed by a trend regression model with change points. The change points are estimated directly from the data. We carry out the analysis for the whole of Germany and the federal state of Bavaria, where we have more detailed data. Both analyses show a major change between March 9th and 13th for the time series of infections: from a strong increase to a decrease. Another change was found between March 25th and March 29th, where the decline intensified. Furthermore, we perform an analysis stratified by age. A main result is a delayed course of the epidemic for the age group 80+ resulting in a turning point at the end of March.

Our results differ from those by other authors as we take into account the reporting delay, which turned out to be time dependent and therefore changes the structure of the epidemic curve compared to the curve of newly reported cases.

## 1 Introduction

The first phase of the Covid-19 pandemic in Germany was managed relatively successfully in comparison to other countries in Europe. Therefore, it is worth taking a closer look at the course of the pandemic in Germany, which has already led to controversial discussions in the public. This particularly concerns the important question about the effectiveness of various control measures. Several publications discuss the effects of control measures in different countries, see, e.g., [1], [2] and [3]. As [4] point out, however, many of such studies are undermined by unreliable data on incidence. Many papers use data provided by the Johns Hopkins University (JHU) [5]. These data are based on cumulative registered cases in different countries, which induces several problems, particularly the fact that not all cases are reported and that there is delay between the day of infection and the reporting day. Furthermore, the systems of reporting vary between countries, which makes comparisons between countries difficult.

In a recent paper on Germany [6], the authors use a complex Bayesian modeling approach based on the daily registrations in the JHU data for Germany. An important claim in [6] is that the lock-down-like measures on March 23rd were necessary to stop exponential growth, however, this result contradicts for example results by the RKI [7]. Furthermore, these approaches were critically questioned by [8] and [9], where the latter emphasized the importance of taking into account the delay by reporting and incubation time, when analyzing the possible effect of non pharmaceutical interventions.

In our analysis we focus on the curve of infection at two geographical levels: the federal state of Bavaria and all of Germany and on four age-strata. The paper is organized as follows. In Section 2, we present the data and the the strategy of estimating the relevant daily counts. Then the segmented regression model, which is the basis for further analyses, is presented. In Section 3, we present the results followed by a discussion in Section 4.

## 2 Data and Methods

### 2.1 Estimation of diseases onset

For the analysis of the Bavarian data, we use the Covid-19 reporting data of the Bavarian State Office for Health and Food Safety (LGL) that is collected within the framework of the German Infection Control Act (IfSG). At case level, this data includes the reporting date (the date at which the case was reported to the LGL) as well as the time of disease onset (here: symptom onset). However, the latter is not always known: partly because it could not be determined and partly because the case did not (yet) have any symptoms at the time of entry into the data base. A procedure for imputation of missing values regarding the disease onset has been developed by [10], using a flexible generalized additive model for location, scale and shape (GAMLSS; [11]), assuming a Weibull-distribution for the time period between disease onset and reporting date. The model includes gender, age, as well as calendar time and day of the week of reporting as covariates. We estimate the delay time distribution from data with disease onset and impute missing disease onsets based on this model.

For the German data, no individual case data were available, so instead we used publicly available aggregated case reporting data published on a daily basis by the RKI [12]. This data contains aggregated numbers of reported cases for all observed combinations of disease onset date and reporting date at local health authorities as well as the numbers of reported cases per day without information on disease onset date. Information is aggregated on district level in different age and sex groups. Based on this aggregated data, we estimated a similar model as described above for the imputation of missing disease onset dates, replacing the smooth associations of age and reporting delay by a categorical effect of the age-group of the cases. To account for differences in reporting behavior in the different federal states, the model was estimated separately for each state and daily onset counts were aggregated after imputation.

We estimated the imputation model for the German data based on all cases reported up until June, 1. The percentage of imputed values was 37% for the Bavarian data and 28% for the German data. Since this percentage is rather high, we performed a sensitivity analysis using (1) only data with a documented disease onset, and (2) utilizing the reporting date of cases as disease onset date when the actual onset date is unknown.

### 2.2 Back-projection

To interpret the course of the epidemic and possible effects of interventions, case based data on time of infection is essential. However, as such data is generally not available, one simple approach is to shift the curve of disease onsets to the past by the average incubation period. The average incubation period for COVID-19 is about five days [13]. A more sophisticated approach, however, is to use the incubation period distribution as part of an inverse convolution, also known as back-projection, in order to estimate the number of infections per day from the time series of disease onsets [14, 15]. We assume a log-normal distribution for the incubation time with a median of 5.1 days and a 97.5% percentile at 11.5 days [13]. These are the same values as used by [6]. For our calculation, we use the back-projection procedure implemented in the R package `surveillance` [16].

### 2.3 The segmented regression model

To analyze the temporal course of the infection we use the following regression model and change points (see [17], [18]):

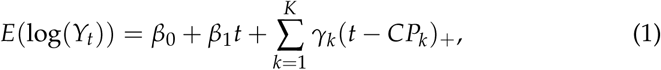

where *Y*_*t*_ is the number of newly infected individuals at time *t, K* is the number of change points, and *x*_+_ = max(*x*, 0) is the positive part of *x*. The change points *CP*_*k*_, *k* = 1, …, *K* are used to partition the epidemic curve *Y*_*t*_ into *K* + 1 phases. These are characterized by different growth parameters. In the phase before the first change point *CP*_1_ the growth is characterized by the parameter *β*_1_, in the 2nd phase between *CP*_1_ and *CP*_2_ by *β*_2_ = *β*_1_ + *γ*_1_. The next change is then at time *CP*_2_. In the 3rd phase between *CP*_2_ and *CP*_3_ the growth parameter is given by *β*_3_ = *β*_1_ + *γ*_1_ + *γ*_2_. This applies accordingly until the last phase after *CP*_*K*_. The quantities exp(*β*_*j*_), *j* = 1, …, *K* + 1 can be interpreted as daily growth factors. Since we use estimated, non-integer values *Y*_*t*_ from the back-projection procedure as outcome, we assume a (conditional) Gaussian distribution for log(*Y*_*t*_). Furthermore, we assume an AR(1) error term for the regression model, since serial correlation occurs due to smoothing in the backprojection procedure.

Since model (1) is a generalized linear model given the change points, the parameters of the model (including the change points) can be estimated by minimizing the respective likelihood function. However, due to the estimation of the change points, the numerical optimization problem is not straight forward. For the estimation of the model we use the R package `segmented`, see [19]. The starting values are partly estimated by discrete optimization. The number of change points *K* is increased from *K* = 1 up to a maximum of *K* = 6. It is examined whether the increase of the number of change points leads to a lower value of the Bayesian information criterion (BIC). Since the considered time series consist of only 61 data points, we exclude models with more than 6 change points, since they are hardly interpretable and the danger of overfitting is high.

We apply the segmented regression model to time series of the estimated daily numbers of infections for Bavaria and Germany. Since the back-projection algorithm yields an estimate for the expected values of the number of daily infections and does so by inducing a smoothing effect, as a sensitivity analysis for the location of the breakpoints, we also apply a regression model to the time series of the daily counts of disease onsets. The results of this sensitivity analysis are presented in the supplementary material. Furthermore, as a more detailed analysis, we apply our procedure to data stratified by age groups. A special focus is on the age group 80+, as this group has the highest risk for a critical course of the disease.

## 3 Results

In Figure 1, the three different time series of daily case counts (newly reported, disease onset and estimated infection date) are presented. The time delay between the three time series for Bavaria and Germany is evident. Furthermore, the curves do not just differ by a constant shift on the x-axis, instead there is also a notable change in the structure of the curves. The curve relating to the date of infection is clearly smoothed due to the back-projection procedure (cf. Section 2.2) and has a clear maximum both for Bavaria and Germany.

**Figure 1:**
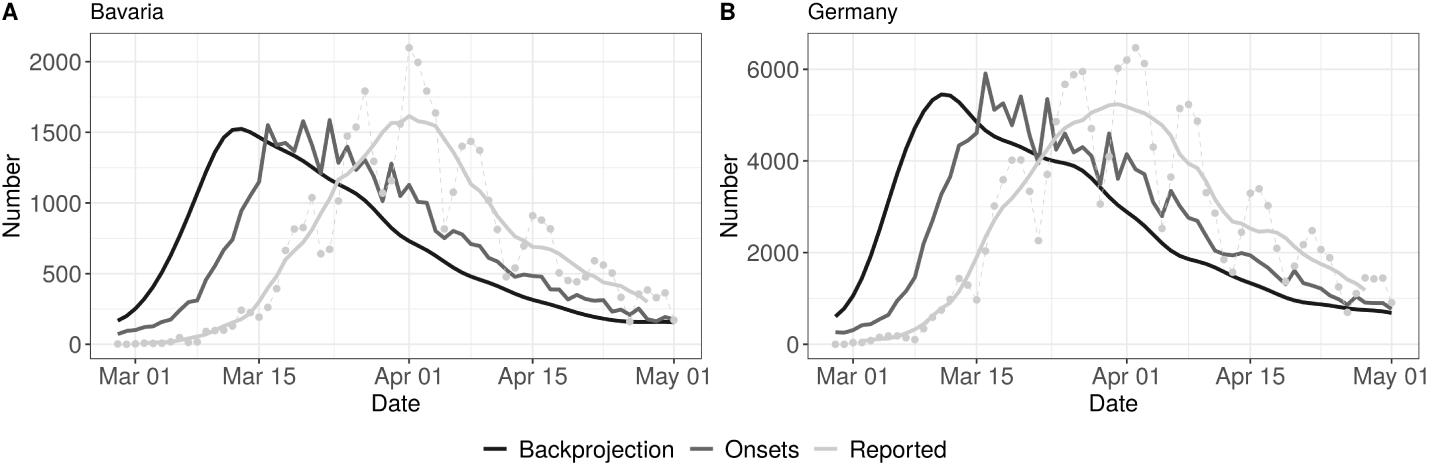
Comparison of time series of daily reported cases (7 day average and daily reported numbers, lightgrey), disease onsets (reported and imputed, grey) and backprojection (derived number of infections, dark grey) for Bavaria (left panel) and Germany (right panel).

### 3.1 Bavarian data

The overall Bavarian model includes five change points. The result can be seen in Figure 2 (left panel) and in Table 1.

**Table 1:**
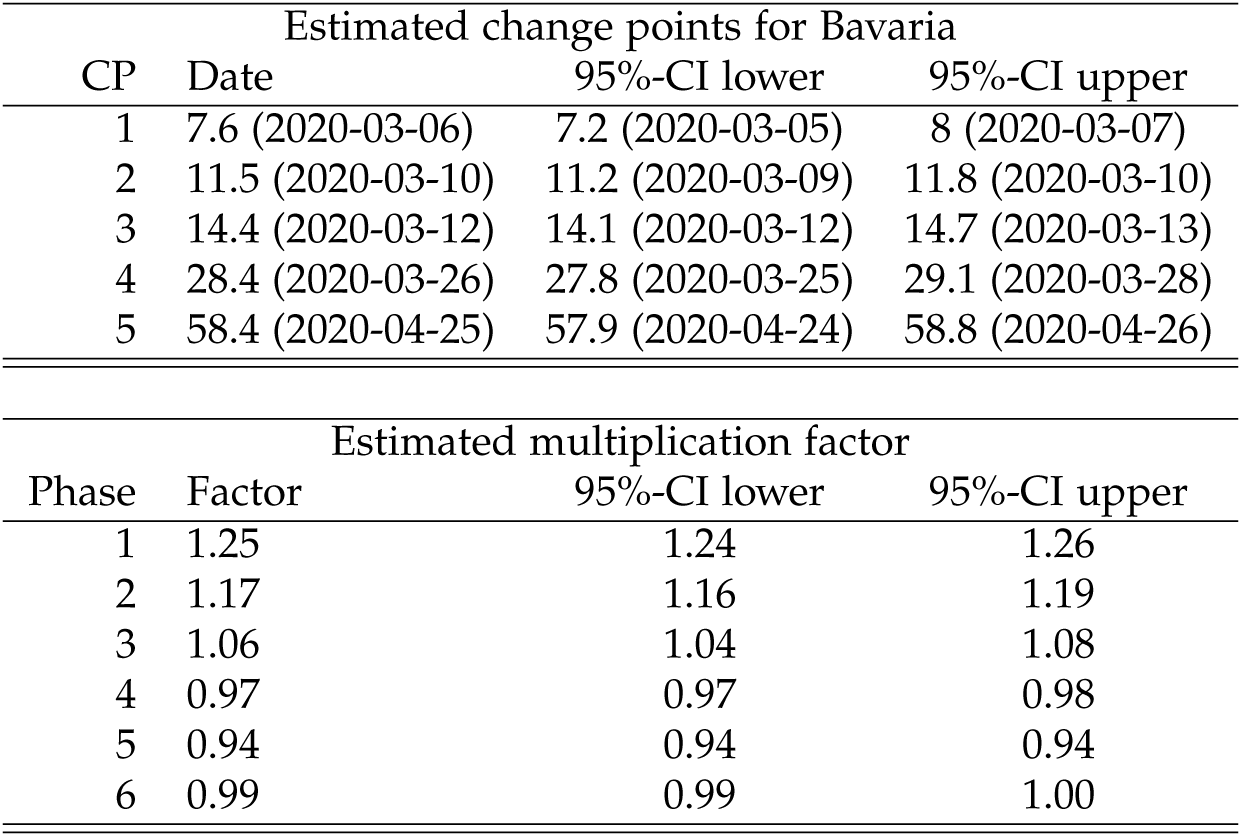
Summary table of the segmented regression model for the expected number of daily infections in Bavaria with five change points. The dates of the estimated change points and the corresponding 95% confidence intervals are given. For the date of the lower/upper limit of the confidence intervals, the values were rounded up or down to the more extreme value. In the second part of the table the estimated multiplication factors of the number of cases per day with the confidence intervals for the 6 phases are given.

**Figure 2:**
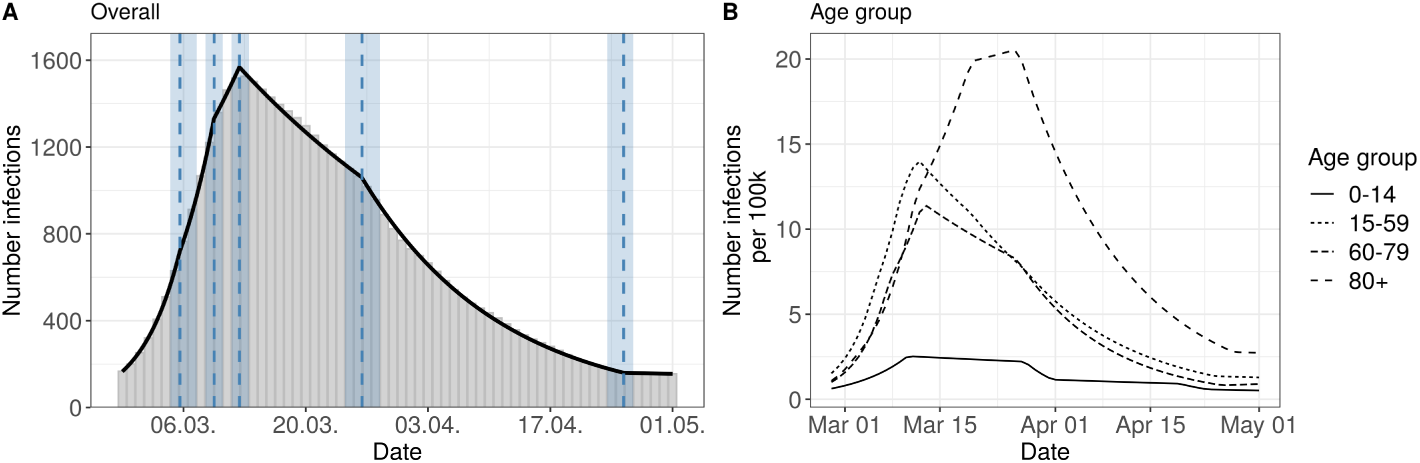
Segmented regression models for Bavaria. The left panel shows results for all reported cases from Bavaria. The solid line is the fitted curve according to the segmented regression model (1) with five change points (*K* = 5) selected based on BIC. The bars are the expected numbers of newly infected cases (cf. Section 2.2). Dashed lines and surrounding shaded ribbons indicate estimated change points and respective, approximate 95% confidence intervals. The right panel shows results of the segmented regression in four age groups based on the back-projected number of infections per 100.000 individuals. The back-projection and segmented regression was estimated in each age-group separately and the selected number of change points varies between groups.

The resulting six phases are:

**1st phase** There is a substantial increase in reported new infections with a high daily multiplication factor of 1.25. The first phase ends at March 6th.

**2nd phase** The increase slows down to a daily multiplication factor of 1.17. This phase lasts to the 10th of March.

**3rd phase** For a short time the multiplication factor further goes down to 1.06

**4th phase** The change point at March 12th marks a clearly visible change in the course of the epidemic. It is the turning point of the curve (change of the multiplication factor to 0.97).

**5th phase** A further change point is found around March 26th. There is an accelerated decrease in the infections with daily factor of 0.94.

**6th phase** On April, 25th the number of infections is rather low and there is no further decrease after this change point (multiplication factor close to 1.)

The age stratified analysis gives some interesting further insights about the course of the epidemic, see Figure 2 (right panel) and Table S1 in the supplementary material. The age groups 15-59y and 60-79y show a similar pattern as the overall analysis. However, in the 80+ age group there are clear differences compared to the other groups. There, the turning point of the epidemic is considerably later on March, 22nd. Furthermore, the number of estimated infections per 100 000 is higher than in the other groups. The age group 0-14y has a much lower number of reported infections, and change points similar to the overall analysis.

### 3.2 German data

The results for the German data are presented in Figure 3 and in Table 2. The model for Germany has four change points inducing five phases:

**Table 2:** Summary table of the segmented regression model for the expected number of daily infections in Germany with four change points. The dates of the estimated change points and the corresponding 95% confidence intervals are given. For the date of the lower/upper limit of the confidence intervals, the values were rounded up or down to the more extreme value. In the second part of the table the estimated multiplication factors of the number of cases per day with the confidence intervals for the 5 phases are given.

| Estimated change points for Germany |  |  |  |
| --- | --- | --- | --- |
| CP | Date | 95%-CI lower | 95%-CI upper |
| 1 | 7.4 (2020-03-05) | 7.2 (2020-03-05) | 7.6 (2020-03-06) |
| 2 | 11.6 (2020-03-10) | 11.3 (2020-03-09) | 11.9 (2020-03-10) |
| 3 | 29.4 (2020-03-27) | 28.4 (2020-03-26) | 30.4 (2020-03-29) |
| 4 | 53.7 (2020-04-21) | 52.4 (2020-04-19) | 55 (2020-04-23) |

Table 2: Summary table of the segmented regression model for the expected number of daily infections in Germany with four change points.
| Estimated multiplication factor |  |  |  |
| --- | --- | --- | --- |
| Phase | Factor | 95%-CI lower | 95%-CI upper |
| 1 | 1.32 | 1.30 | 1.33 |
| 2 | 1.11 | 1.09 | 1.13 |
| 3 | 0.98 | 0.97 | 0.98 |
| 4 | 0.94 | 0.94 | 0.95 |
| 5 | 0.97 | 0.96 | 0.98 |

**Figure 3:**
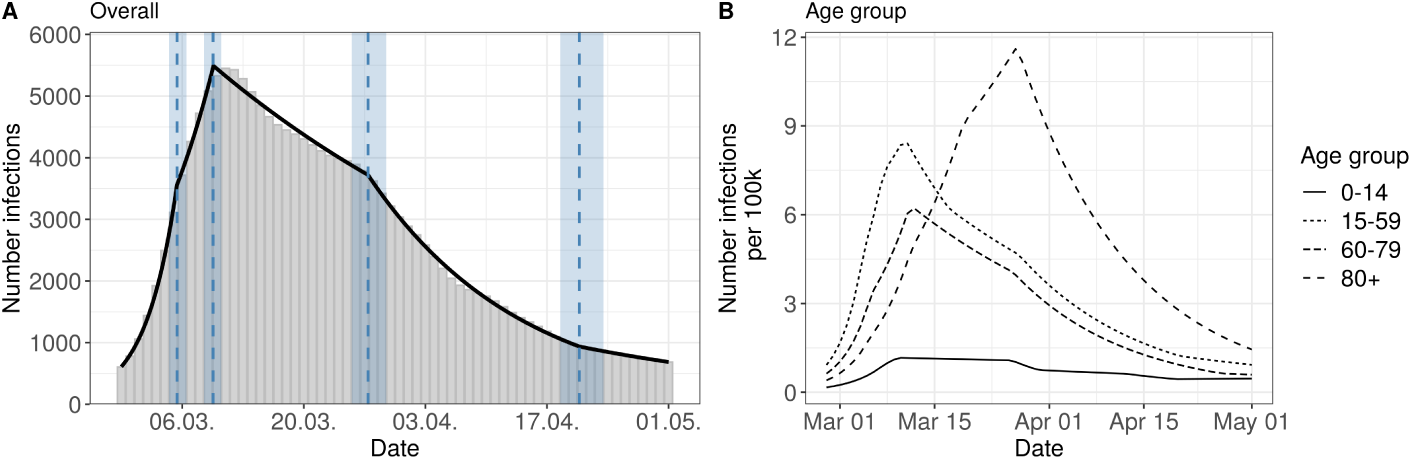
Segmented regression models for Germany. The left panel shows results for all reported cases. The solid line is the fitted curve according to the segmented regression model (1) with six change points (*K* = 6) selected based on BIC. The bars are the expected numbers of newly infected cases (cf. Section 2.2). Dashed lines and surrounding shaded ribbons indicate estimated change points and respective, approximate 95% confidence intervals. The right panel shows results of the segmented regression in four age groups based on the back-projected number of infections per 100.000 individuals. The back-projection and segmented regression was estimated in each age-group separately and the selected number of change points varies between groups.

**1st phase** There is a strong increase (multiplication factor 1.32 per day) in new infections in the beginning of the epidemic until March 5th, where the increase decreases.

**2nd phase** From March 5th to March 10th there is still a substantial increase of infections with a daily multiplication factor around 1.1.

**3rd phase** After the change point at March 10th, there is a clearly visible change in the course of the epidemic. There is change from an increasing to a decreasing curve with a daily multiplication factor of about 0.98. This phase lasts until 27th of March.

**4th phase** An acceleration of the decrease can be seen from March 27th onward.The daily multiplication factor is 0.94

**5th phase** After April, 21st, the number of infections is rather low with a slow decrease.

The results of the age stratified analysis are similar to the results for Bavaria (cf. Figure 3, right panel and Supplement Table S2). The age group 80+ once again differs substantially from the other age groups. The turning point for the 80+ group is on March 25th. The other age groups show a structure similar to the overall analysis.

## 4 Discussion

The present analysis is a retrospective, exploratory analysis of the German and Bavarian COVID-19 reporting data during Mar-Apr 2020.

### 4.1 Limitations

The analysis only includes reported cases. If the proportion of undetected cases changes over time, e.g., due to different test criterion, this can distort the curve and thus the determination of the change points. Therefore, additional data on daily deaths and hospital admissions and the number of tests performed should be considered. Furthermore, it is possible to estimate the proportion of undetected cases with the help of representative studies such as the one currently conducted in Munich, see [20]. In a recent paper [21] performed an time varying estimation of the case detection ratio (CDR) for different age groups. They find a linear decreasing CDR from March 2nd until April 12th in the main age groups. The CDR was only half as large at the end of the period as it was at the beginning. This can only partly explain the curve, where we observe much a much higher increase.

Our analysis is based to a considerable extent on imputed data, see [10], which is a results of missing data w.r.t. the disease onset. We have performed a sensitivity analysis using only cases with available disease onset date and based on imputing missing disease onset dates by the reporting date of the cases (Figure S2 in the supplementary material).

The back-projection procedure is based on an assumption of the distribution of the incubation time. There are some recent papers showing some evidence for a longer incubation time in elderly cases, see [22] and [23]. Therefore, we performed an additional sensitivity analysis comparing the results for the 80+ age group for different assumptions about the incubation time distribution, see supplementary material Figure S3. We find in this sensitivity analysis that the curve of the new infections is shifted to the left by about 2-3 days. However, the structure of the curve does not change considerably. Altogether, the much later peak in the 80+ compared to the other population strata cannot only be due to a different incubation time.

Since changes in behavior do not occur abruptly, the assumption of change points is also problematic in itself. Therefore, the interpretation of change points should always be done in conjunction with a direct observation of the epidemic curve.

### 4.2 Interpretation of Results

Our analysis is based on the onset of the disease (more precisely: the onset of symptoms) and a back-projection to the date of infections, and therefore, despite its limitations, is better suited to describe the course of the epidemic than the more common analysis of daily or cumulative reported case numbers.

In the analysis of the Bavarian and the German data our main result is the change point, where the exponential growth was stopped: this clearly happened already between March 9th and 13th. The timing of this change point coincides to the implementation of the first control measures: the partial ban of mass events with more than 1000 people. Furthermore, in a press conference on March 11th chancellor Merkel and the president of the RKI appealed to self-enforced social distancing [24]. Furthermore, the extended media coverage from Bergamo, Italy, as well as the voluntary transition to home-office work could be related to this essential change in the course of the pandemic.

In Bavaria and in Germany, the change point at 26/27th of March of infection date is apparent. This change point is associated with different measures taken in March (closing of schools and stores on March 16th and the shutdown including contact ban on March 22nd in Germany including Bavaria). Other measures were similar in timing in Bavaria and all over Germany. In Bavaria, some measures were implemented a little earlier. Since there were many measures administered simultaneously and since – as described above – other factors beyond the measures itself contributed, we do not think is not possible to quantify the effect of individual measures to the development of the epidemic curve.

The results for the 80+ age group indicate a infection curve which is delayed by about one week compared to the other age groups. This is possibly due to the fact that the disease was first introduced into Germany by younger holidaymakers and business travelers. Hence, it likely took some additional generations of transmission before the infections mitigated into the 80+ group. Furthermore, many infections in the age group 80+ are due to outbreaks in nursing homes and homes for the elderly, where very different mechanisms of contact occur compared to the rest of the population. Many of the restrictions were targeted at the younger age-groups (school closings, mobility restrictions), hence, the effect of these interventions is only indirect for the 80+ group and thus the impact is delayed. This underlines the need for more direct measures for this group. For the age group of 0-14 the breakpoints are similar to the other age groups. The much lower infection rate could be partly due to lower case detection ratio, since infected children show less symptoms, see e.g. [25].

The claim by Dehning et al. [6], that the shutdown on March 21st was necessary to stop the growth of the epidemic is not supported by our analysis. There is a change point in the epidemic curve after that date, but the major change from an exponential growth to a decrease was before the shutdown. The difference in results can be explained by the different data bases used for the respective analyses. While Dehning et al. [6] use data based on daily registered cases, in our analysis, data on disease onset are included. As can be seen from Figure 1 and from the results of our data analysis, the delay distribution of the time between disease onset and reporting day changed over time. This makes a crucial difference. In a recent technical addendum [26] the authors re-fit their model on more appropriate data. These analysis – in our opinion – clearly show that the effective reproduction number decreased earlier than in their initial analysis, however, they attribute the decrease to a SIR model peculiarity, where a linear decrease in the contact rate can lead to the incidence curve dropping despite *R*(*t*) *>* 1.

The above discussions illustrate how complex the interpretation of even simple SIR models is and the question is, if such SIR modeling is not too simple to really allow for questions to be answered model based (no age structure, no time varying reporting delay, no incubation period). In contrast, our approach is more data driven with a minimum of modeling assumptions and without the need to include strong prior information about the change points. Directly using a segmented curve with exponential growth (decline) is in line with common models of infectious diseases in its early stages, where the limitation of the spread by immune persons plays no role. The problem of using complex models with many parameters for the evaluation of governmental measures has also been highlighted by [8].

Our approach is similar to that of [9] who performs a change point analysis for the cumulative reported numbers as well as the estimated *R*(*t*). The use of the time-varying reproduction number *R*(*t*), a standard measure to describe the course of an epidemic is challenging, as different definitions have been proposed in the literature that also imply different interpretations (see [27, 28]). However, the analysis of R(t) as a relative measure can be useful, when one wants to analyze data from different countries with non comparable reporting systems, see [3].

Altogether, the effect of governmental measures as whole is clearly documented in the literature, see, e.g., [1] and [2]. Our results are in line with that of [29], where a stop of exponential growth in Great Britain was also observed before the lockdown. Our result on a possible effect of the ban of mass events is also in line with the results of [3].

The temporal connection between the change points in our analysis and various control measures should be interpreted as an association, rather than a direct causal relationship. In the end many other explanations exists and from a simple time series analysis it is not possible to say to what extent the population already had changed their behavior voluntarily, as for example observed in mobility data [30, 31], and in what way the measures contributed to this. More speculative alternative explanations would include the possibility of a seasonal effect on coronavirus activity (e.g. related to temperature) or changes in test capacity or the case detection ratio. However, given the re-emergence of the epidemic in the fall of 2020 at high test capacity and at relatively high temperatures shows that contact behavior is the major explanatory factor for virus activity. Nevertheless, any analysis of observational time series data including only a limited amount of explanatory factors has to be interpreted with care and with respect to the many uncertainties which remain regarding COVID-19 [32].

Despite the limitations of the approach, we argue that it is advantageous and important to directly interpret the epidemic curve and the absolute number of cases, rather than indirect measures like the *R*(*t*). Furthermore, the reproduction rate does not contain information about how many people are currently affected, or whether the infected persons belong to risk groups. The course of the time-varying reproduction number calculated by us for Bavaria fits well with the change point analysis [10]. A value of *R*(*t*) >1 corresponds to a rate of increase >1, noting that the time delays in the interpretation of *R*(*t*) must be kept in mind.

It should be noted, that the presented analysis is retrospective. Control measures have to be decided based on a completely different level of information than what the retrospectively established epidemic curve suggests. The simple observation of the course of the reported case numbers by reporting date is also problematic because this course is strongly influenced by the reporting behavior and the methods and capacities of the test laboratories. Typically, substantially fewer cases are reported at weekends than during the week. Therefore, the estimation proposed by Guenther et al. [10] is an important step to estimate the better interpretable curve of new cases, but is limited by assumptions and limitations itself, that need to be considered when interpreting the results.

Since the impact of the measures also depends on how they are implemented by the population (compliance), the results cannot be directly transferred to the future. Nevertheless, it remains a remarkable result that the clear turning point of the early COVID-19 infection data in Germany is associated with non drastic measures (no shutdown) and strong appeals by politicians.

### Data and code availability

Data used for the analyses and all code to reproduce the models, figures and tables in the manuscript are openly and freely available from [33]. All analyses were performed using the R programming language [34].

## Supporting information

Supplementary Analyses

## Data Availability

Data used for the analyses and all code to reproduce the models, figures and tables in the manuscript are openly and freely available.

https://zenodo.org/record/4449816

## Acknowledgements

We would like to thank Katharina Katz and Manfred Wildner from the Bavarian State Office for Health and Food Safety (LGL) for providing the data and for useful discussions. We also thank Nadja Sauter for help with visualizations. We thank two reviewers whose comments helped to improve the work significantly.

