## Supplementary Analyses for "Analysis of the early Covid-19 epidemic curve in Germany by regression models with change points"

### Supplemental Material

#### Segmented regression model for disease onsets

We use the model

$$E(Y_t) = \exp\left(\beta_0 + \beta_1 t + \sum_{k=1}^{K} \gamma_k (t - CP_k)_+\right), \tag{1}$$

where $Y_t$ is the number of new disease onsets at time $t$, $K$ is the number of change points, and $x_+ = \max(x, 0)$ is the positive part of $x$.

We apply a negative binomial model as flexible model for the counts. The model is fitted by maximum likelihood and the BIC information criterion is used to assess the number of change points. This is done by discrete optimization followed by applying the R-Package *segmented*, see Muggeo (2008). The results for German and Bavarian date are presented in table S1 and in figure S1.

**A** Bavaria

| Estimated change points | | | |
|---|---|---|---|
| CP | Date | 95%-CI lower | 95%-CI upper |
| 1 | 18 (2020-03-16) | 17.9 (2020-03-15) | 18.1 (2020-03-17) |
| 2 | 32.4 (2020-03-30) | 32.2 (2020-03-30) | 32.7 (2020-03-31) |

| Estimated multiplication factor | | | |
|---|---|---|---|
| Phase | Factor | 95%-CI lower | 95%-CI upper |
| 1 | 1.20 | 1.20 | 1.20 |
| 2 | 0.98 | 0.98 | 0.98 |
| 3 | 0.94 | 0.94 | 0.94 |

**B** Germany

| Estimated change points | | | |
|---|---|---|---|
| CP | Date | 95%-CI lower | 95%-CI upper |
| 1 | 6.5 (2020-03-04) | 6.3 (2020-03-04) | 6.6 (2020-03-05) |
| 2 | 12.5 (2020-03-11) | 12.4 (2020-03-10) | 12.7 (2020-03-11) |
| 3 | 18 (2020-03-16) | 17.8 (2020-03-15) | 18.2 (2020-03-17) |
| 4 | 34.7 (2020-04-02) | 34.3 (2020-04-01) | 35 (2020-04-02) |

| Estimated multiplication factor | | | |
|---|---|---|---|
| Phase | Factor | 95%-CI lower | 95%-CI upper |
| 1 | 1.17 | 1.17 | 1.18 |
| 2 | 1.33 | 1.32 | 1.33 |
| 3 | 1.10 | 1.09 | 1.11 |
| 4 | 0.98 | 0.98 | 0.98 |
| 5 | 0.95 | 0.95 | 0.95 |

Table S1: Summary table of the segmented regression model for the number of disease onsets in Bavaria and Germany with two and four change points selected based on BIC. The dates of the estimated change points and the corresponding 95% confidence intervals are given. For the date of the lower/upper limit of the confidence intervals, the values were rounded up or down to the more extreme value. In the second part of the table the estimated multiplication factors of the number of cases per day with the confidence intervals for the 3/5 phases are given.

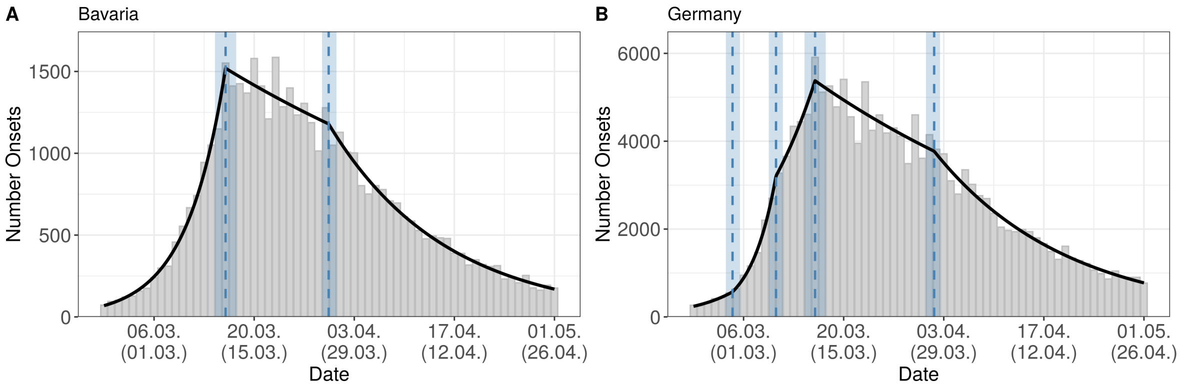


Figure S1: Segmented regression models for Bavaria and Germany based on the daily number of disease onsets. The left panel shows result for the selected model with two change points for the Bavarian data. The right panel shows the selected model with four change points for Germany. The solid line is the fitted curve according to the segmented negative binomial regression model. The bars are the (observed and imputed) numbers of daily disease onsets. Dashed lines and surrounding shaded ribbons indicate estimated change points and respective, approximate 95% confidence intervals. The x-axis labels correspond to the day of disease onset in the first row, in the second row we show the approximate day of infection in brackets (day of disease onset -5 days).

### Sensitivity analysis for imputation of missing onset dates

We show results of sensitivity analyses for two changes in the imputation of missing disease onset dates. In the first sensitivity analysis, we focus only on cases with available disease onset date, in the second analysis, we set the onset date to the reporting date for all cases without information on the disease onset date. The results are presented in FigureS2. When focusing on cases with available onset dates only (sens 1), the absolute number of infections per day are smaller, but the general structure of the curve is very similar to the main analysis. When utilizing reporting date as onset date for individuals without onset information the decline of infections in the second half of March is less strong but still observable. The peak of the number of infections is identical for all three curves.

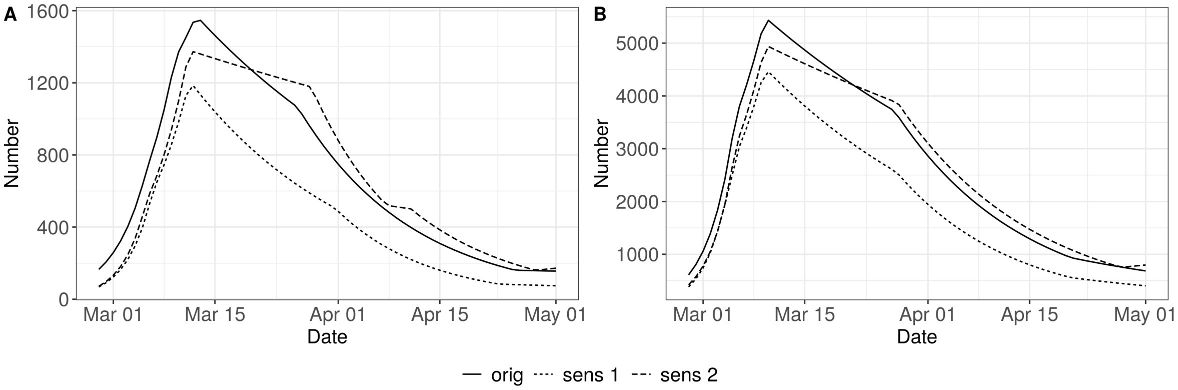


Figure S2: Segmented regression models for Bavaria and Germany based on the backprojected number of infections. Shown are the results with imputed disease onsets (original analysis), based on cases with reported disease onset dates only (sens 1), and based on all cases where missing disease onset dates are set to the reporting date of the respective case (sens 2) for Bavaria (left panel) and Germany (right panel).

### Detailed results from the age stratified analysis

| | Age group | | | |
|---|---|---|---|---|
| CP | 0-14 | 15-59 | 60-79 | 80+ |
| 1 | 03-10 [03-09, 03-11] | 03-06 [03-05, 03-06] | 03-08 [03-07, 03-08] | 03-02 [03-01, 03-03] |
| 2 | 03-27 [03-26, 03-29] | 03-09 [03-08, 03-10] | 03-12 [03-12, 03-13] | 03-11 [03-10, 03-12] |
| 3 | 04-01 [03-30, 04-02] | 03-12 [03-11, 03-12] | 03-26 [03-25, 03-28] | 03-20 [03-18, 03-21] |
| 4 | 04-19 [04-18, 04-21] | 03-20 [03-17, 03-22] | 04-26 [04-25, 04-27] | 03-26 [03-25, 03-28] |
| 5 | 04-23 [04-22, 04-25] | 03-27 [03-25, 03-30] | | 04-27 [04-26, 04-29] |
| 6 | | 04-25 [04-23, 04-26] | | |
| Phase | 0-14 | 15-59 | 60-79 | 80+ |
| 1 | 1.13 [1.11, 1.15] | 1.26 [1.25, 1.27] | 1.26 [1.24, 1.27] | 1.27 [1.24, 1.30] |
| 2 | 0.99 [0.98, 1.01] | 1.16 [1.15, 1.18] | 1.11 [1.08, 1.13] | 1.19 [1.18, 1.21] |
| 3 | 0.85 [0.81, 0.89] | 1.07 [1.04, 1.10] | 0.98 [0.97, 0.98] | 1.06 [1.05, 1.08] |
| 4 | 0.99 [0.98, 1.00] | 0.97 [0.96, 0.98] | 0.93 [0.92, 0.93] | 1.01 [0.99, 1.02] |
| 5 | 0.90 [0.85, 0.94] | 0.96 [0.95, 0.96] | 1.02 [1.00, 1.03] | 0.94 [0.93, 0.94] |
| 6 | 0.98 [0.96, 1.01] | 0.94 [0.94, 0.94] | | 1.00 [0.97, 1.02] |
| 7 | | 0.99 [0.98, 1.00] | | |

Table S2: Age-group specific results for Bavarian data. The upper panel shows the estimated change points with 95%-confidence intervals. The lower panel shows the estimated daily multiplication factors with 95%-confidence intervals.

| | Age group | | | |
|---|---|---|---|---|
| CP | 0-14 | 15-59 | 60-79 | 80+ |
| 1 | 03-08 [03-07, 03-08] | 03-04 [03-04, 03-05] | 03-06 [03-05, 03-06] | 03-05 [03-04, 03-07] |
| 2 | 03-10 [03-09, 03-10] | 03-07 [03-07, 03-08] | 03-11 [03-11, 03-12] | 03-12 [03-10, 03-13] |
| 3 | 03-26 [03-25, 03-27] | 03-11 [03-10, 03-11] | 03-27 [03-25, 03-28] | 03-19 [03-18, 03-21] |
| 4 | 03-30 [03-29, 03-31] | 03-17 [03-15, 03-19] | 04-27 [04-25, 04-28] | 03-27 [03-26, 03-28] |
| 5 | 04-12 [04-11, 04-14] | 03-27 [03-26, 03-29] | | |
| 6 | 04-20 [04-18, 04-21] | 04-20 [04-19, 04-22] | | |
| Phase | 0-14 | 15-59 | 60-79 | 80+ |
| 1 | 1.23 [1.21, 1.24] | 1.35 [1.34, 1.36] | 1.29 [1.27, 1.30] | 1.27 [1.25, 1.29] |
| 2 | 1.12 [1.08, 1.16] | 1.16 [1.14, 1.19] | 1.12 [1.10, 1.13] | 1.17 [1.15, 1.19] |
| 3 | 1.00 [0.99, 1.00] | 1.05 [1.03, 1.07] | 0.97 [0.97, 0.98] | 1.09 [1.07, 1.10] |
| 4 | 0.91 [0.90, 0.93] | 0.95 [0.94, 0.96] | 0.94 [0.94, 0.94] | 1.03 [1.02, 1.05] |
| 5 | 0.99 [0.98, 0.99] | 0.97 [0.97, 0.98] | 0.98 [0.97, 1.00] | 0.94 [0.94, 0.95] |
| 6 | 0.95 [0.94, 0.97] | 0.95 [0.94, 0.95] | | |
| 7 | 1.00 [1.00, 1.01] | 0.97 [0.97, 0.98] | | |

Table S3: Age-group specific results for German data. The upper panel shows the estimated change points with 95%-confidence intervals. The lower panel shows the estimated daily multiplication factors with 95%-confidence intervals.

### Sensitivity analysis for back-projection in the age-group 80+

We compare two scenarios for the backprojection in the age group 80y+. In the original analysis, we assume a log-normal distribution with median 5.1 and 97.5%-quantile 11.5 days ($LN(1.63, 0.415)$). As a sensitivity analysis we use a log-normal distribution with median incubation time of 7.7 and 95%-quantile 14.1 days (97.5%-quantile 15.83, $LN(2.04, 0.368)$). The resulting curves differ by a slight shift but they are identical in structure.

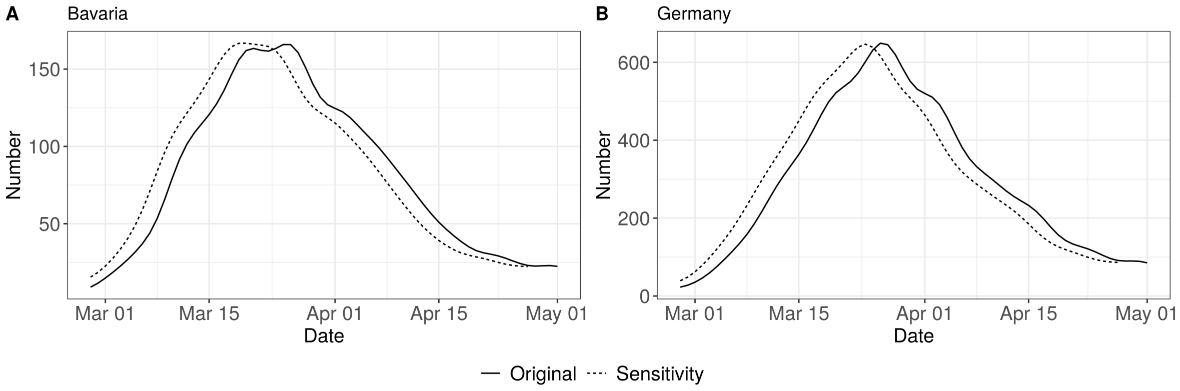


Figure S3: Expected numbers of new infections per day for the age group 80+ in Bavaria and Germany under two assumptions regarding the incubation time distribution.
